# Fine-scale variation in the effect of national border on COVID-19 spread: A case study of the Saxon-Czech border region

**DOI:** 10.1101/2022.03.01.22271644

**Authors:** Adam Mertel, Jiří Vyskočil, Lennart Schüler, Weronika Schlechte-Wełnicz, Justin M. Calabrese

**Affiliations:** Center for Advanced Systems Understanding (CASUS), Görlitz, Germany; Department of Computational Hydrosystems, Helmholtz Centre for Environmental Research (UFZ), Leipzig, Germany; Helmholtz-Zentrum Dresden Rossendorf (HZDR), Dresden, Germany; Department of Ecological Modelling, Helmholtz Centre for Environmental Research (UFZ), Leipzig, Germany; Department of Biology, University of Maryland, College Park, MD, USA

## Abstract

The global extent and temporally asynchronous pattern of COVID-19 spread have repeatedly highlighted the role of international borders in the fight against the pandemic. Additionally, the deluge of high resolution, spatially referenced epidemiological data generated by the pandemic provides new opportunities to study disease transmission at heretofore inaccessible scales. Existing studies of cross-border infection fluxes, for both COVID-19 and other diseases, have largely focused on characterizing overall border effects. Here, we couple fine-scale incidence data with localized regression models to quantify spatial variation in the inhibitory effect of an international border. We take as a case study the border region between the German state of Saxony and the neighboring regions in northwestern Czechia, where municipality-level COVID-19 incidence data are available on both sides of the border. Consistent with past studies, we find an overall inhibitory effect of the border, but with a clear asymmetry, where the inhibitory effect is stronger from Saxony to Czechia than vice versa. Furthermore, we identify marked spatial variation along the border in the degree to which disease spread was inhibited. In particular, the area around Löbau in Saxony appears to have been a hotspot for cross-border disease transmission. The ability to identify infection flux hotspots along international borders may help to tailor monitoring programs and response measures to more effectively limit disease spread.

## Introduction

The COVID-19 pandemic is an event of unprecedented magnitude in modern world history and has consequently set off a flurry of research activity across the health-related sciences. These efforts have generated an equally unprecedented deluge of spatially referenced epidemiological data. High resolution epidemiological data have the potential to reveal disease transmission processes on heretofore inaccessible spatial scales, but significant challenges in data harmonization and modeling remain. The global extent and temporally asynchronous nature of the COVID-19 spread have repeatedly put the spotlight on international borders and the policies governing them. Research focusing on cross-border infection fluxes can thus deepen our understanding of COVID-19 spread dynamics while also serving to inform border-related policy decisions.

Several papers have studied COVID-19 border dynamics for different regions with a range of methodologies. For example, Grimmé et al. (2021) used the Endemic-Epidemic framework (Bekker-Nielsen Dunbar and Held, 2020) to model the effect of policies on Swiss-Italian borders. Eckardt, Kappner, and Wolf (2020) applied a Bayesian spatio-temporal Poisson model to study the role of border controls in the Schengen Area, and Hossain et al. (2020) used a metapopulation model for estimating the effect of travel restrictions on the COVID-19 outbreak. Additionally, Laroze et al. (2021) constructed a multi-source spatial model on top of the pre-crisis commute to work to study the effects of borders between regions within the same country, and Han et al. (2021) explored the balance between domestic and imported cases in Chinese cities. While all of these studies suggested a significant effect of border presence and border control regime on the spread of COVID-19, all focused on determining the overall effect of the focal borders.

Most of the border-focused studies mentioned in the previous paragraph are based on the spatial level of whole countries, regions, or spatially separated metropolitan areas. More generally, most spatio-temporal COVID-19 models are applied either on the level of whole countries or on medium-sized provinces and regions due to data limitations. In contrast, we developed a model of COVID-19 spread on the very detailed level of individual municipalities. While several studies consider this level of granularity for data analysis and modeling (e.g., Arauzo-Carod, 2021; Cole et al., 2020; Neyens et al., 2020; Schuler et al., 2021), research combining datasets on this scale from more than one country is still exceedingly rare even it may provide an unique value for studying the spatiotemporal trends in detail. Furthermore, while governing policy may be uniform across an entire border, variation in geomorphology, population distribution, transportation infrastructure, socio-economical aspects, history and other factors along a border may generate spatial variation in the inhibitory effect of the border on disease spread. To our knowledge, this type of fine-scale variation has not been studied in the context of COVID-19 spread. We therefore aim to quantify cross-border infection fluxes at the scale of individual municipalities in the Saxon-Czech border region.

Our approach is based on the simple assumption that the virus travels mainly with humans. Therefore, COVID-19 may be transmitted more easily between two localities with a higher intensity of mobility connecting them than between localities with lower mobility. Several approaches are widely used in epidemiology to estimate mobility. Probably the most common is the gravity model, which has many possible adaptations and extensions (Yan and Zhou, 2019) and has been a mainstay of spatial COVID-19 modeling (e.g., Chen et al., 2021; Ezzat et al., 2021; Werner, 2021; Zhu et al., 2021).

The basic gravity model estimates the mobility between two places primarily from two variables - their separation distance and the population size of both places. The gravity model can be further extended to include border effects. This idea was applied in the context of various topics, e.g., international to domestic trade (McCallum, 1995), the cross-border flows of immigration (Lewer and Van den Berg, 2008), air traffic (Becker et al., 2018), or mergers and acquisitions (Wong, 2008). Even more relevant to this study is the paper of Kramer et al. (2016), in which the authors extended the gravity model with a border effect to study the international transmission of Ebola virus.

Here, we propose a set of local multiple regressions–inspired by the principles of the gravity model–that allow us to quantify fine-scale spatial variation in the effect of the border. Consistent with other studies, we find that the Saxon-Czech border has an overall inhibitory effect on COVID-19 spread. However, we also identify both between-country asymmetry and substantial fine-scale variation in the strength of the border effect. In particular, the border in the region around Löbau in Saxony appears to have had a weaker inhibitory effect than other areas along the Saxon-Czech border. We conclude by discussing how high resolution border studies can potentially pinpoint hotspots for disease spread where additional border monitoring and controls may be warranted.

## Data

For Saxony, we used the dataset “Gemeindegrenzen 2018 mit Einwohnerzahl” (GeoBasis-DE et al., 2020) for the geometries and population sizes on the municipality (“*Gemeinde”*) level. Czech population numbers on the municipality level (“obec”) were taken from the Czech Statistical Agency (Czech Statistical Office, 2021), while the geometries were obtained from RÚIAN (Czech Office for Surveying, Mapping and Cadastre, 2021). We further joined the population size numbers with their associated geometries and merged the datasets from both countries. To keep the same geometry detail on both sides of the borders, we applied the Douglas-Peucker (Douglas and Peucker, 1973) simplification algorithm implemented in the Python library topojson (mattijn, 2021).

The number of infections in single municipalities in Saxony is published by the coronavirus.sachsen.de (Sächsische Staatsregierung, 2021). Unfortunately, the website does not provide any historical data in a tabular format, just a table with data from the last seven days. We therefore set up a CRON job to visit the website every day and scrape the contents in the form of a .csv table. On the Czech side, we worked with the daily updated dataset from the Czech Ministry of Health (Komenda et al., 2021). We also reclassified both datasets to weekly granularity to avoid problems with weekly cyclicity (i.e., pronounced decreases in cases reported on weekends, national holidays etc.). For this study, we used only data for the three regions in Czechia that share a border with Saxony - Liberec, Ústí nad Labem, and Karlovy Vary. The entire region defined for the case study is shown in Figure 1.

**Figure 1.**
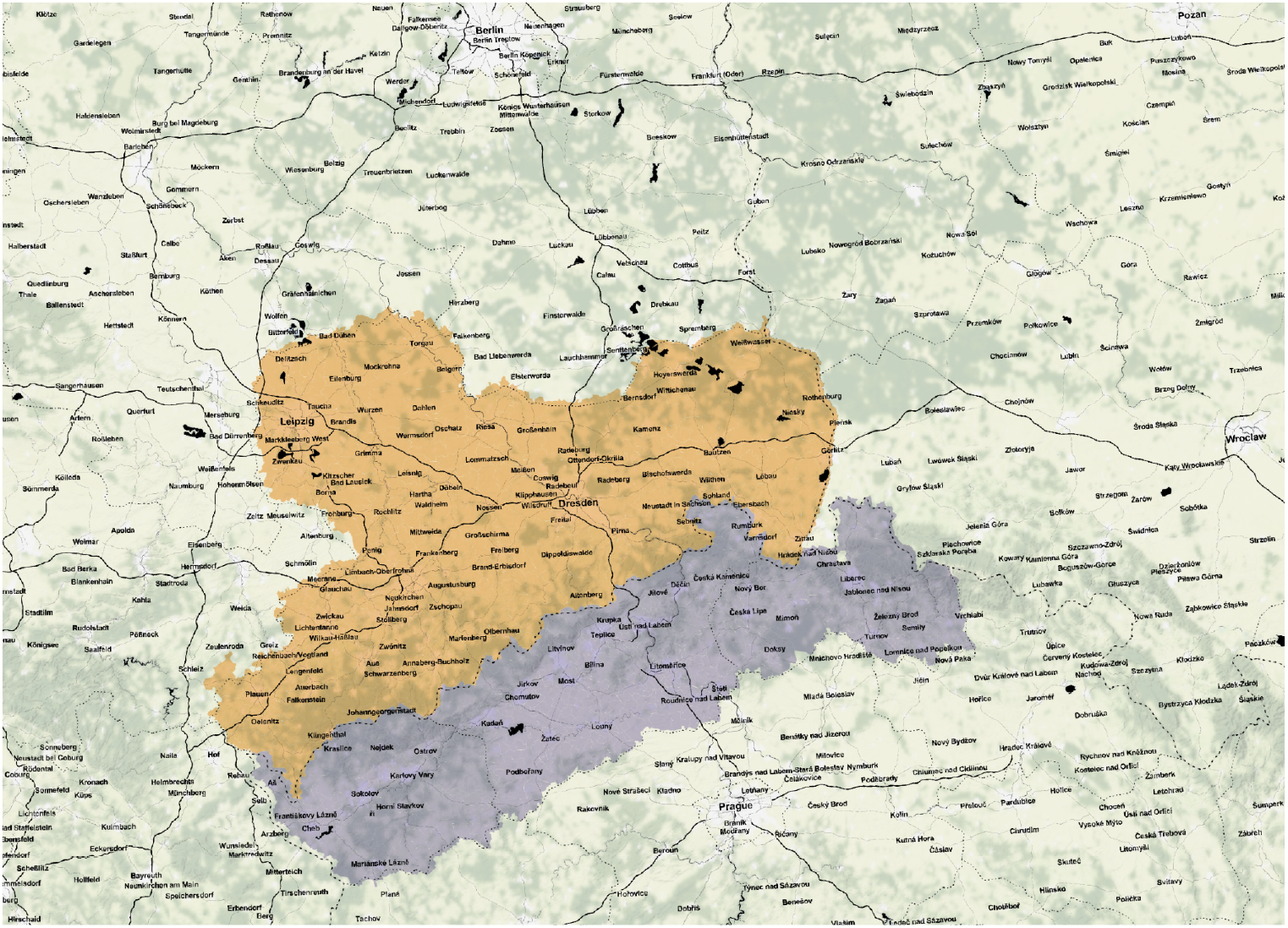
Overview map shows the location of the case study area and the extent of two selected bordering regions - the federal state Saxony in Germany, and the regions of Liberec, Ústí nad Labem and Karlovy Vary in Czechia.

After data cleaning and harmonization, we obtained a dataset of 1116 municipalities (almost 6 million inhabitants), with 415 located in Saxony (4,077,937 inhabitants), and 701 in Czechia (1,810,283 inhabitants). The municipalities in Czechia are smaller on average, which reflects a slightly different administrative organization. The timeframe of the case study ranges from the 31st of January to the 13th of June 2021. This timeframe was selected due to the availability of the data on both sides of the border, but also to capture the wave of high case numbers in late winter and spring 2021.

Most of the above-cited spatial epidemiology papers use the Euclidian distance between location centroids as the measure of distance. However, this method is problematic on the granular spatial scale of our study. Instead, the connectivity between municipalities is mostly defined by the infrastructure that is determined by social (e.g., culture, work/school commuting, local subsidies) and physical (e.g., mountainous areas, rivers) factors. To account for variation in these structural factors, we estimate “temporal distances” between pairs of municipalities via the typical car-based driving time between them.

To calculate the temporal distances between municipalities, we firstly needed to extract the point representing the real central place of each municipality. This was done by the search engine Nominativ (osm-search, 2021), which offers a free API service to geocode places by their names. For every request, it returns the point coordinates that represent the geographical unit. For municipalities, the API returns mostly the point of the central square or the centroid of the built-up areas. This point was then used as a starting point for the calculation of isochrones. For this step, we used the Openroute API (GIScience Research Group and HeiGIT, 2021) that returned the areas accessible by car within a specific time limit. We set the upper value to 1 hour and worked with 10 six minute intervals. Then, the temporal distance was calculated by iterating through all pairs of municipalities and checking into which isochrone interval of the first municipality the centroid of the second municipality falls. This measure of distance between two municipalities accounts for the transportation networks connecting them and is thus more informative for our purposes than the simple Euclidean distance.

We performed several exploratory analyses before implementing our regression models (see Figures S1, S2, S3, and S4). Here, we quantified the overall patterns in the temporal trends, the spatial co-occurrence of the case number increases, and the effect of the spatial and temporal autocorrelation. We conclude that the pandemic situation in each country developed somewhat independently, with indications of a significant but heterogeneous spatial effect of the national border. Case numbers in municipalities within a short driving distance (12 minutes and less) are much more strongly correlated than those separated by longer distances. Additionally, municipalities’ case numbers are, in general, more correlated at shorter time lags, with the lowest variance in correlations occurring at time lags of 1 to 2 weeks.

## Model

To quantify the local effect of the national border on the spread of COVID-19, we constructed a set of beta regression models, with one for each municipality. Inspired by the gravity model, each local model quantifies the contributions of neighboring municipalities to an index of “virus import potential” for the focal municipality as a function of population sizes, inter-municipality temporal distances, and presence/absence of the border between each pair of municipalities. We calculated virus import potential by comparing the increases of cases in each focal municipality with the pandemic situation in other municipalities in its neighborhood.

The computation of the model starts by iterating through the list of all municipalities, and for each focal municipality *a*, we define a neighborhood of municipalities *{b, c*, …, *f}*, which are reachable from *a* in less than forty minutes of driving time in a car. This value is supported by additional travel data for this geographical region (Bundesagentur für Arbeit, 2020; Centrum dopravního výzkumu, v. v. i., 2019; infas Institut für angewandte and Sozialwissenschaft GmbH, 2017). These sources show that the largest distance people usually travel to work, school, or shopping is not more than 20 kilometers / 30 minutes. But there are types of travel that often occur over longer distances and times, such as tourism, family visits, and business-related journeys that may take even more than one hour / 100 kilometers. We therefore chose forty minutes as a conservative value that would comfortably accommodate most normal travel.

For each municipality *a*, we constructed a local beta regression model that was based on the characteristics of the relations of *a* to the municipalities in the neighborhood *{b, c*, …, *f}*. As a dependent value for the model, we calculated the possibility of importing the virus from *{b, c*, …, *f}* to *a*. First, we identified all the weeks when the number of cases in *a* was rising. We focused on the weeks when the number of cases in the municipality was increasing because he spatial covariances may differ in various periods of the pandemic (Schuler et al., 2021) in this model. For each of the increasing weeks *t*, we looked at the situation in the neighborhood to identify the places, where the increase may have been spread from. We looked at the number of cases within the municipalities in the neighborhood of *{b, c*, …, *f}* at *t*_*-1*_ that is one week before the increase in *a*. Then, we relativized the numbers for *{b, c*, …, *f}* to the sum of all cases in the neighborhood, so we ended with virus import potential values scaled to [0, 1]. We repeated this process for every week *t* in the dataset. Finally, we calculated mean values for all combinations of municipalities in the dataset, and then transformed the values from [0, 1] to (0, 1) so that they could be used as the dependent variable in a beta (multiple) regression model. This calculation does not directly measure the real import of the virus. Instead, it is a reasonable approximation of the possibility to explain an increase in municipality *a* with the number of cases in municipality *b*, one week before this increase. We chose the lag of one week based on the common understanding of the virus spread and the time-lagged correlation analysis (Figure S4).

We added three predictor variables to the model. The first of them was closeness, which was calculated as the inverse value of the relative temporal distance. The higher the closeness value, the faster it was possible to reach the focal municipality from the second municipality by car. Second, we used each municipality’s population size as a proxy for its attractivity. The third predictor variable was the presence of the border. Here, the value 1 was assigned to the municipalities in the neighborhood that were not located in the same country. For example, when *a* was in Saxony, we assigned 0 to all the Saxonian municipalities and 1 to all Czech municipalities, and vice versa when *a* was in Czechia.

The linear predictor of the mean of our beta regression model, which is linked to the (0, 1) index of virus import via the logit link function, can thus be written as

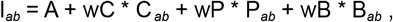

where *I*_*ab*_ stands for the virus import potential from municipality *b* to municipality a, and A is an intercept. *C*, P, and *B* are the closeness of municipality *a* and *b*, the relative size of municipality *b* compared to the maximal municipality population size within the neighborhood of municipality *a*, and the existence of borders between municipality *a* and *b*, respectively. *wC, wP*, and *wB* represent the regression coefficients of these parameters. The spatial pattern of *wB* is of particular interest to us. Specifically, spatial variation in this coefficient allows us to quantify and visualize spatial variation in the border effect. We then modeled the precision parameter of the beta regression as a linear function (identity link) of border presence, wB_Φ_ * B_*ab*_, which improved both model fit and convergence relative to a constant precision model.

For each municipality *a*, we calculated a model consisting of the dependent variable and all three predictor variables for each municipality in the neighborhood *{b, c*, …, *f}* for the mean of the response, and a separate model consisting of just the border term for the precision of the response. We first standardized predictors of relative municipality size and closeness variables via z-scores, and then estimated the beta models via maximum likelihood. These analyses were implemented in R (R Core Team, 2000) and the betareg package (Zeileis et al., 2016). For a visual guide of the whole procedure, please refer to Figure 2.

**Figure 2.**
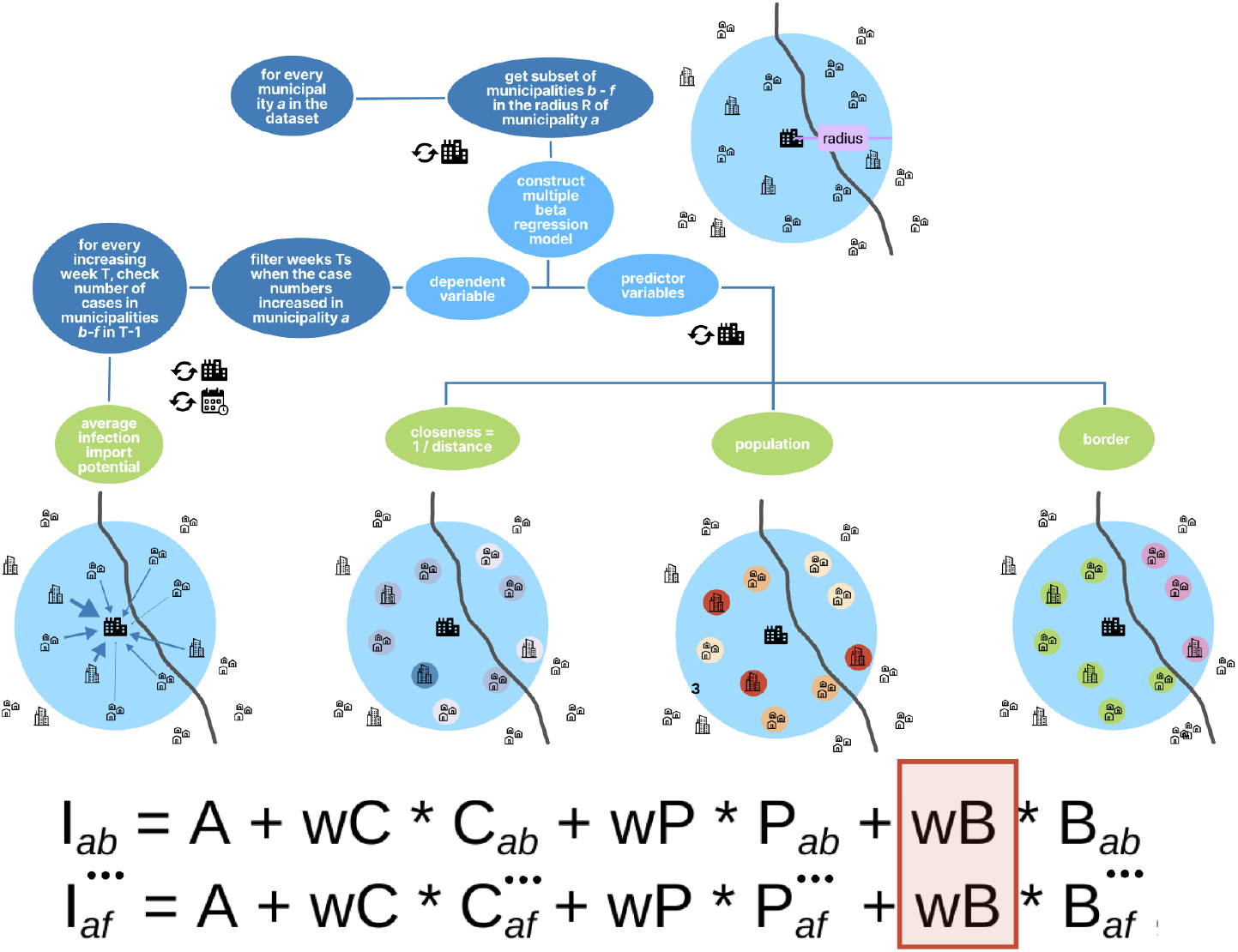
Visual overview of the model construction framework.

To validate the results of the model, we did two post-hoc analyses. The first check is based on a comparison of the average of all weeks when cases in the focal municipality increased weighted by the magnitudes of the increases (referred to as central increase week further in the study). This number represents the differential timing of the waves in the two countries as an alternative way to look at the degree of cross-border coupling. The second post-analysis to validate the results of the model is based on the idea of the Granger causality metric (Granger, 1969; Romero García et al., 2021), which tests the causation of one time series by another time series considering a time lag. For each municipality *m*, we constructed two microregions. The first one consisted of the municipalities falling into a driving distance limit of 20 minutes (half of the threshold used for the model) from *m* and the same country as *m*, the second consisted of the municipalities in the driving distance of 40 minutes from *m* but located on the other side of the border. We summed weekly case numbers within both microregions and calculated the symmetrized changes, which tend to be more robust compared to classical percentage change (Berry and Ayers, 2006). These values were further used to perform the Granger causality test and the correlation to the weighted average of the border effect value (weighted by the population sizes of the municipality).

## Results

The model coefficients (*wD, wB*, and *wP*) are estimated based on the three predictor variables for each municipality, from which the *wB*, the effect of the border, was the focal point of our study. The spatial distribution of this value is displayed on map in Figure 3. This map shows that within most of the municipalities in our study area, the border strongly inhibits virus spread (pink to magenta colors), but with pronounced the spatial variation in the strength of this inhibitory effect. In particular, the national border appears to have a less inhibitory effect (greener colors) in the area around the town of Löbau in eastern Saxony.

**Figure 3.**
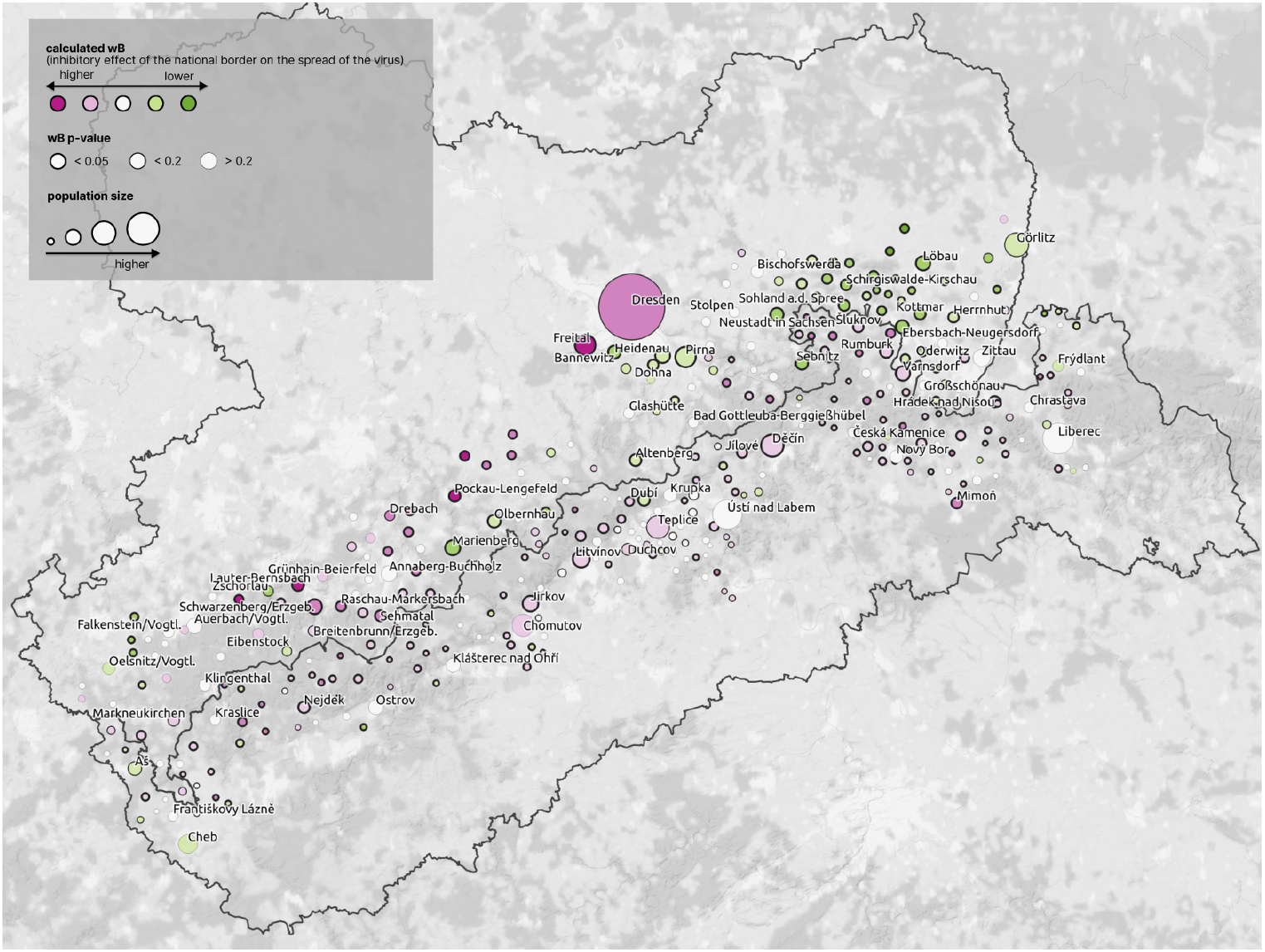
Map of municipalities in the neighborhood of the Saxon-Czech border. The circle size represents the population, and the color depicts the estimated *wB*, which represents the potential of the border to inhibit the spread of COVID-19. The green color stands for a less inhibitory effect of the border, while magenta tones represent municipalities where the border strongly inhibited disease spread. The width of the circle stroke represents the p-value of the border parameter of the municipality.

In total, 365 local models were constructed, one for each municipality that fulfilled the condition to have at least 5 municipalities in the neighborhood that lay on the other side of the border. Only one of those models did not converge, and was therefore removed from the analysis. Across all the remaining 364 local models, the median r^2^ value is ∼0.37, and the median p-value is ∼0.05. Our methodology was not capable of identifying statistically significant results (p < 0.05) mostly for the municipalities located on the outskirts of the study area, which may have been influenced by regions not considered in this study. Results also highlight marked asymmetry on opposite sides of the national border, with a higher potential of virus import from Czechia to Saxony than vice versa (visible also on Figure S1 and Figure S2). This may be explained by the overall spatio-temporal trend of the spread over the timeframe of our study, in which the Northern Czech Republic (and specifically the province of Cheb) was reported as a European hotspot at the very beginning of the year 2021, followed by high incidence values in Saxony in the following months (March, April).

The border region near the towns of Löbau and Bautzen in eastern Saxony stands out as a hotspot where increases in focal municipalities are more strongly linked to case number increases on the Czech side of the border. There are also single municipalities in the other parts of both Saxony and Czechia that show weaker inhibition (e.g., Marienberg, Johanngeorgenstadt, Neuhausen, and Dubí located in the central part of the study area). Outside of these exceptions, the overall effect of the border on disease transmission is mixed or strongly inhibitory.

The spatial distribution of central increase week is shown on the left map of Figure 4. In this picture, we can identify the spatial discontinuity created by the national border. This effect is least pronounced at two places - around the town of Šluknov (including the German neighborhoods of Löbau and Sebnitz) where the COVID-19 wave came later (during mid-March) compared to other Czech regions, and the westernmost part of Saxony, where the case numbers peaked relatively early compared to the rest of Saxony. The right map then shows the neighborhood variance (within 40 minutes of driving distance) of this value. Because the central increase week values form, in general, two spatial clusters - one for each country, the places with high values of the variance are then concentrated around the national border. One exception of this trend is located around the towns of Löbau (Germany) and Šluknov (Czechia), which again indicates a less inhibitory effect of the national border on the virus spread in this area. This result is consistent with the outcome of the beta regression analysis, which shows the highest potential for cross-border virus import exactly in this area.

**Figure 4.**
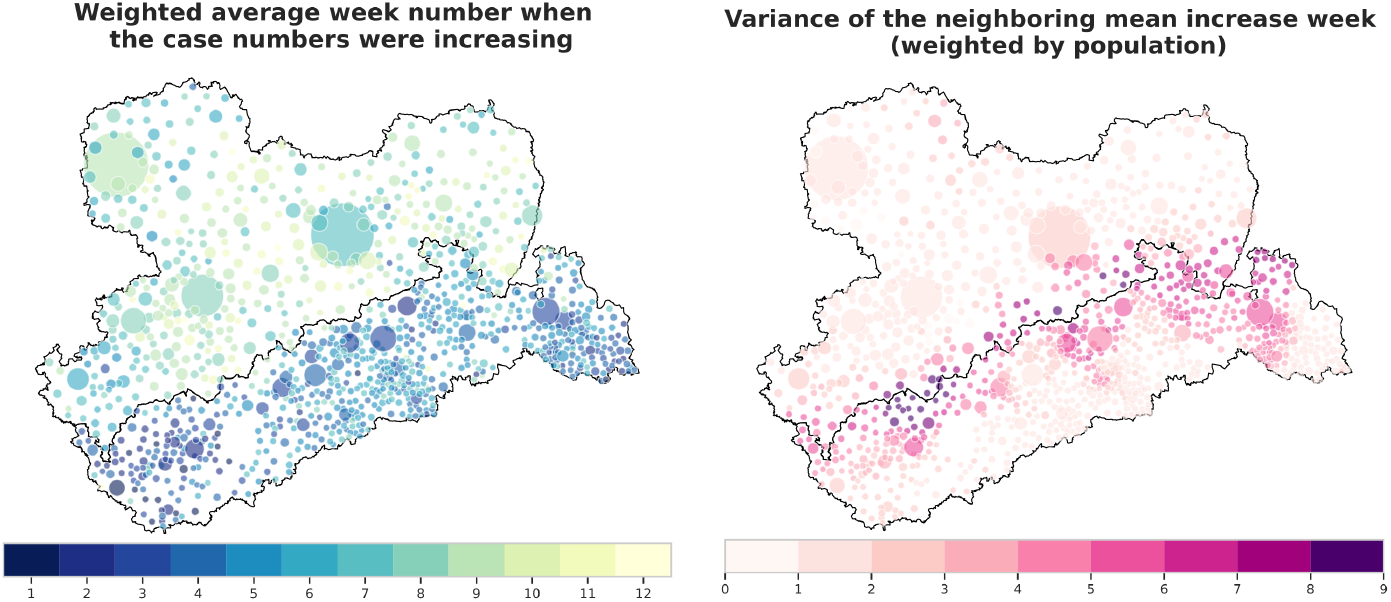
Weighted central increase week (left) and the neighborhood variance in central increase week (right).

Figure S5 shows the comparison of Granger causality p-values for the time lags of one, two, and three weeks vs. the weighted average border effect value (weighted by the population sizes within the microregion) that estimated via the beta regression models. It shows that the weaker coefficient of the border effect, the more significant the Granger causality tended to be. The correlation coefficients are -0.4 in the case of the maximal time lag of 1 week, -0.46 for the maximal lag of two weeks, and -0.54 for the maximal lag of three weeks.

## Discussion

In this paper, we proposed a regression-based methodology to estimate spatial variation in the effect of a national border on disease spread. Our approach leverages fine-scale, spatially-referenced case count data to estimate a location-specific index of virus import potential. We then demonstrated the power of our techniques on a municipality-level case study of COVID-19 spread in the Saxon-Czech border region. Consistent with other studies of border effects, we found an overall inhibitory effect of the national border on disease spread between the two regions, but with stronger inhibition in the direction of Saxony to Czechia than vice versa. Importantly, our approach also highlighted pronounced spatial variation in the inhibitory effect of the border, particularly in the region around Löbau in Saxony. Specifically, the border provided much weaker inhibition of disease spread in this region from Czechia to Saxony compared to most other areas along the border. This hotspot was clearly identified by our local regression models, and this central result was also verified by post-hoc analysis focusing on differences in the timing of the outbreaks on either side of the border.

In the hotspot around Löbau, the timing of the localized case count peak was much closer to that of the neighboring region in Czechia (neighborhood of town Šluknov) than it was to other accessible municipalities in Saxony. This finding suggests a pronounced, cross-border coupling of epidemic dynamics in this region, which our regression models clearly identified. In contrast, the central region of the border displayed a stronger inhibitory effect, with the disease dynamics between neighboring regions of Saxony and Czechia more clearly decoupled. With respect to the strength of the border effect, the western-most areas near towns of Plaunen and Cheb were intermediate relative to the eastern and central border regions.

We suspect that these localized differences in border effect, particularly those between the eastern (Löbau) and central border regions, are driven by differential cross-border traffic patterns. Even though the borders were “closed” for most of the time span covered by our case study, there were several exceptions valid for crossing in one or both directions. First of all, commuters traveling for work from the Czech Republic to Saxony, on a daily or weekly basis, are essential to several sectors of the Saxonian economy, including healthcare, tourism, transport, and manufacturing (Sujata et al., 2020). Some of these workers had exceptions even during the hardest lockdown, some were pushed to minimize border crossings (*Sachsen*.*de*, 2021a; *Sachsen*.*de*, 2021b). The state authorities announced mandatory testing in January 2021 for all cross-border workers (*Sachsen*.*de*, 2021c), which increased the case numbers on the German side. On the other hand, people from Saxony used to go shopping in Czechia regularly due to the lower prices of some commodities (e.g., gas, groceries, and some services), and the closure of the border would sharply reduce such traffic. Physical conditions also modulate cross-border mobility. For example, the Ore Mountains (Erzgebirge / Krušné Hory) along the central part of the Saxon-Czech border form a natural barrier for many cross-border activities. The distribution of cities and areas of higher population density between sides of the borders can also have an effect. Specifically, the central region of the border features larger cities on the Czech side than on the German side. When coupled with the mountainous terrain along the border and sparser road networks, this difference in population density may reduce border crossings both for Czechs (less need to commute for work) and for Germans (less convenient to commute for commodities, leisure). In contrast, the opposite factors would likely contribute to increased border traffic in the region around Löbau. While our results are consistent with this hypothesis, explicit data on cross-border movements, which were unavailable to us, would be required to confirm this interpretation fully.

A limitation of our study is the lack of direct consideration of some of the surrounding regions, which may have influenced COVID-19 dynamics near the Saxon-Czech border, notably Poland to the far east and the German states of Bavaria and Thuringia to the west. The unavailability of municipality-scale data from these regions precluded their consideration in our analysis. This issue manifests itself in the lower pseudo *R*^*2*^ values for the models and higher *p*-values on the border term that tend to occur on the outer edges of our study area. This is specifically relevant for the regions of Görlitz in the east and Cheb / Plauen in the west. Outside of these outlying regions, we are confident that our analysis accurately quantifies disease import potential from surrounding regions and provides reliable results.

This robustness is further evidenced by the agreement between our core model-based analysis and the confirmatory analyses based on timing differences and on Granger causality.

Our index of virus import is based on a comparison of the case increase in a focal municipality with the relative number of cases in the neighboring municipalities one week earlier. Calculation of this index therefore requires determining appropriate values for neighborhood size, time lag, and relative weightings of contributions with the neighborhood. While our selection of these parameters was guided by literature review and knowledge gained via exploratory analysis, we tested the robustness of our results by various driving distance thresholds, temporal lags, and weighting techniques. Similarly, we considered a range of options for dealing with exact 0’s and 1’s in our index of virus import. Collectively, these alternative modeling assumptions do slightly affect our numerical results; however, these variations changed neither our qualitative results nor the inferences we drew from our analysis. Finally, we urge caution in interpreting patterns in our index of virus import. While this metric identifies areas of the higher potential for cross-border disease transfer, it does not directly measure such transfer. In other words, the high estimated potential for cross-border transmission does not guarantee that such transmission actually happened.

We have developed a method for quantifying fine-scale variation in the effect of a border on between-country (or between-region) disease transmission potential. A key advantage of our approach is that it does not require data on cross-border movements of individuals, which are generally difficult to obtain and come with substantial privacy concerns. Furthermore, our approach is able to detect potential hotspots for cross-border disease transmission, which might then be flagged for additional monitoring efforts or stricter border measures in the future. While this type of analysis is retrospective, it could potentially be automated such that, as new data become available, it provides near real-time results on patterns of cross-border disease transmission. Such an automated system could help authorities decrease the time needed to adjust border policies to changing epidemic conditions. A related issue concerns determining the extent to which such cross-border transmission hot spots are consistent over time. The multiple waves of COVID-19 that have occurred worldwide should provide ample opportunities to address this and related questions in future studies.

## Data Availability

All data produced in the present study are available upon reasonable request to the authors

## Acknowledgments

This work was partially funded by the Center of Advanced Systems Understanding (CASUS), which is financed by Germany’s Federal Ministry of Education and Research (BMBF) and by the Saxon Ministry for Science, Culture, and Tourism (SMWK) with tax funds on the basis of the budget approved by the Saxon State Parliament.

## Supplementary Material

**Figure S1.**
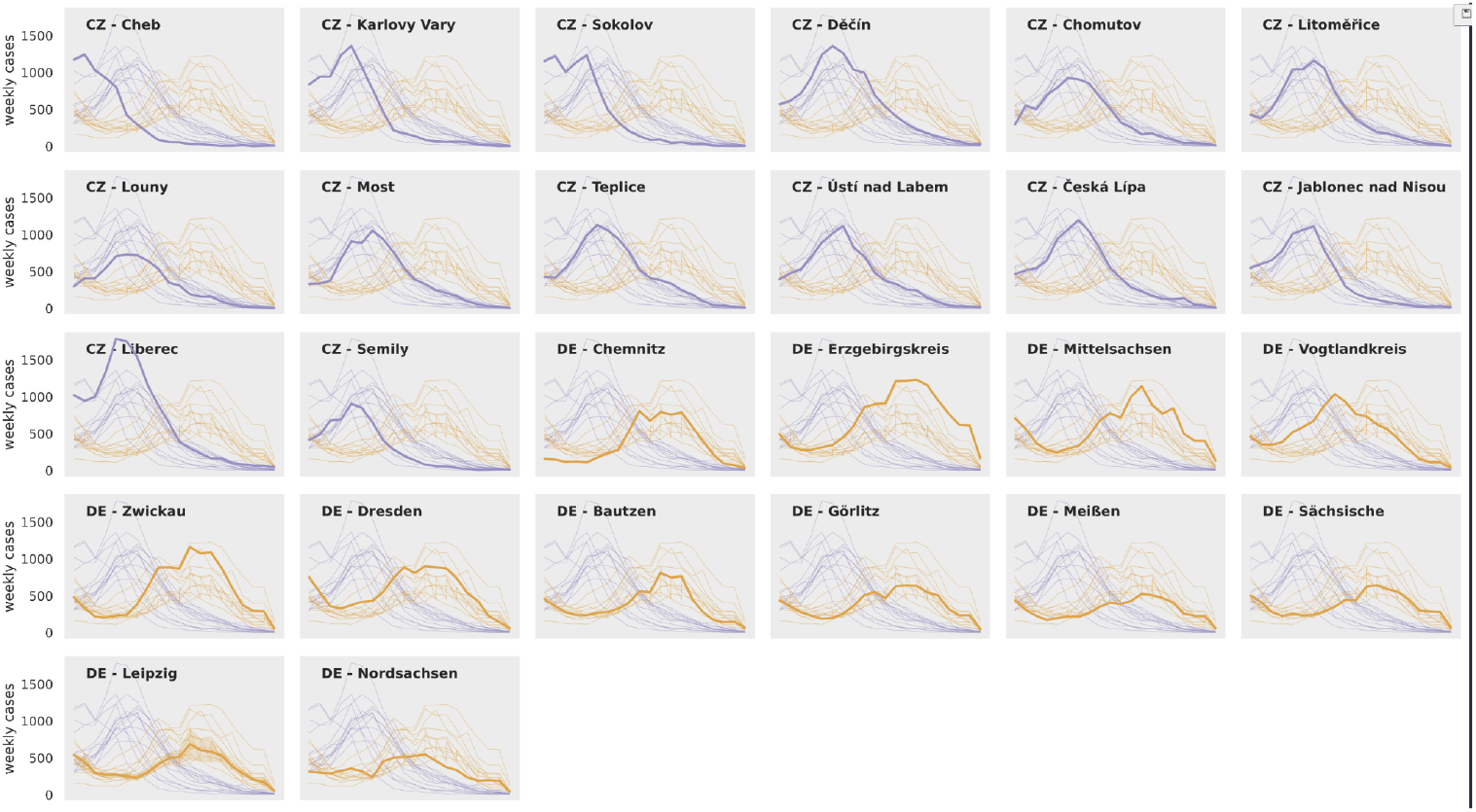
Temporal patterns of the weekly cases in every region (14 “okres” in CZ and 12 “Kreise” in DE) show a substantial difference between the trends of the German and Czech regions. While the numbers in the Czech regions tend to rise at the beginning of the timeframe (February), German regions have a wave of high numbers in the middle of the timeframe (late March, April). Even though the trends within each country are similar, considerable differences also exist among neighboring regions. Although the Czech regions are considerably smaller in population, the number of cases is similar, implying a higher incidence on the Czech side.

**Figure S2.**
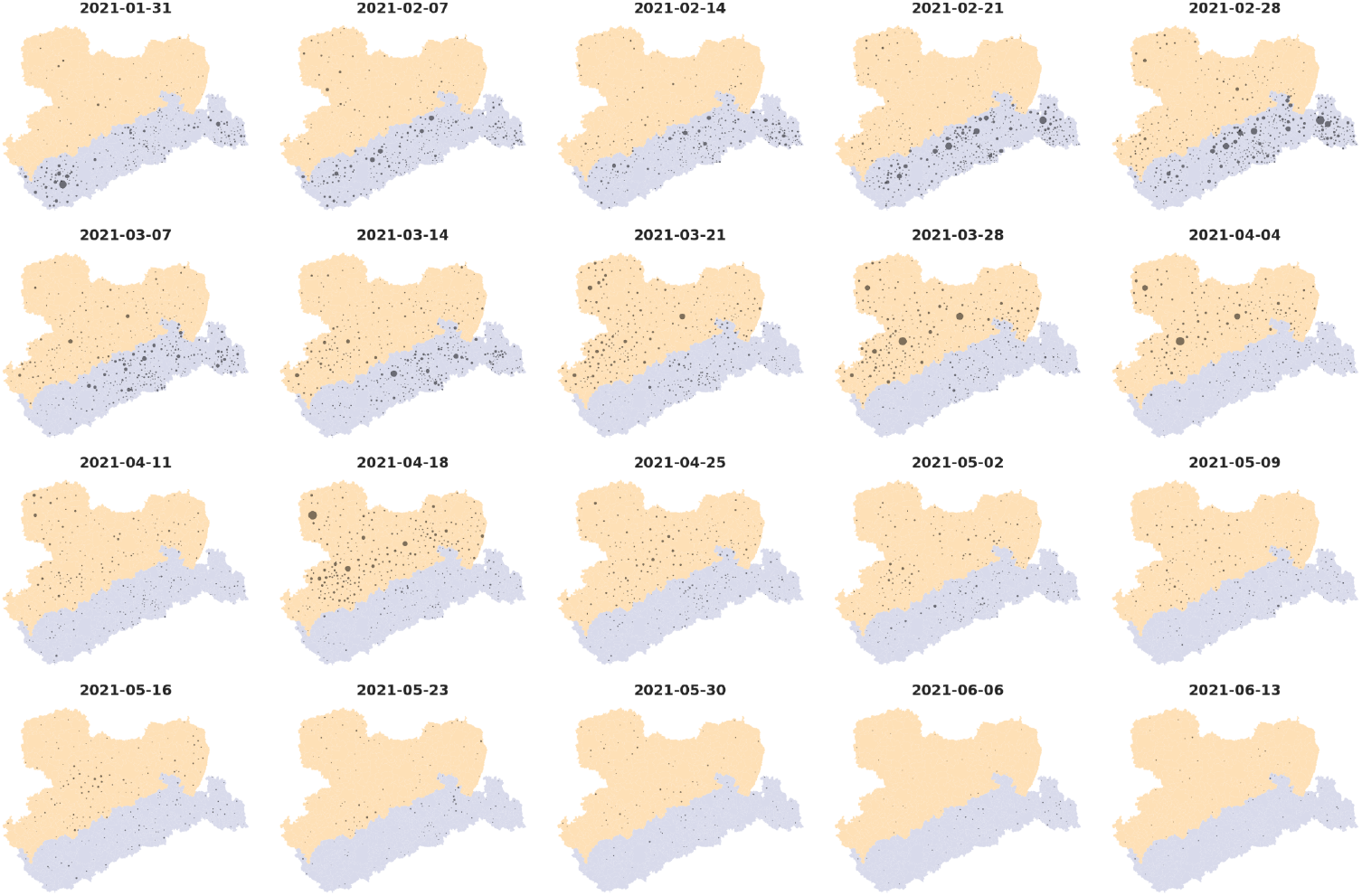
Weekly increases in the number of cases on the level of municipalities. This figure shows the municipalities with increased cases each week of the time series. Here we can follow the general spatio-temporal trends of the spread. At the very beginning of the timeframe, the situation was considerably worse in the western part of Czechia, particularly around the towns of Karlovy Vary and Cheb. During February, the wave of high numbers came to all Czech regions, while from March, the places with increases in case numbers are more dispersed and mostly located within the regions around Děčín, Ústí nad Labem, and Chomutov. On the German side, the first more significant and spatially concentrated wave of increased cases appeared in the middle of March in the region of western Saxony, around Plauen and Zwickau. The following week, we can see an increase also in north-western Saxony (Leipzig region), followed by the central (Dresden, Chemnitz) and then eastern (Görlitz, Bautzen, Zittau)regions of Saxony.

**Figure S3.**
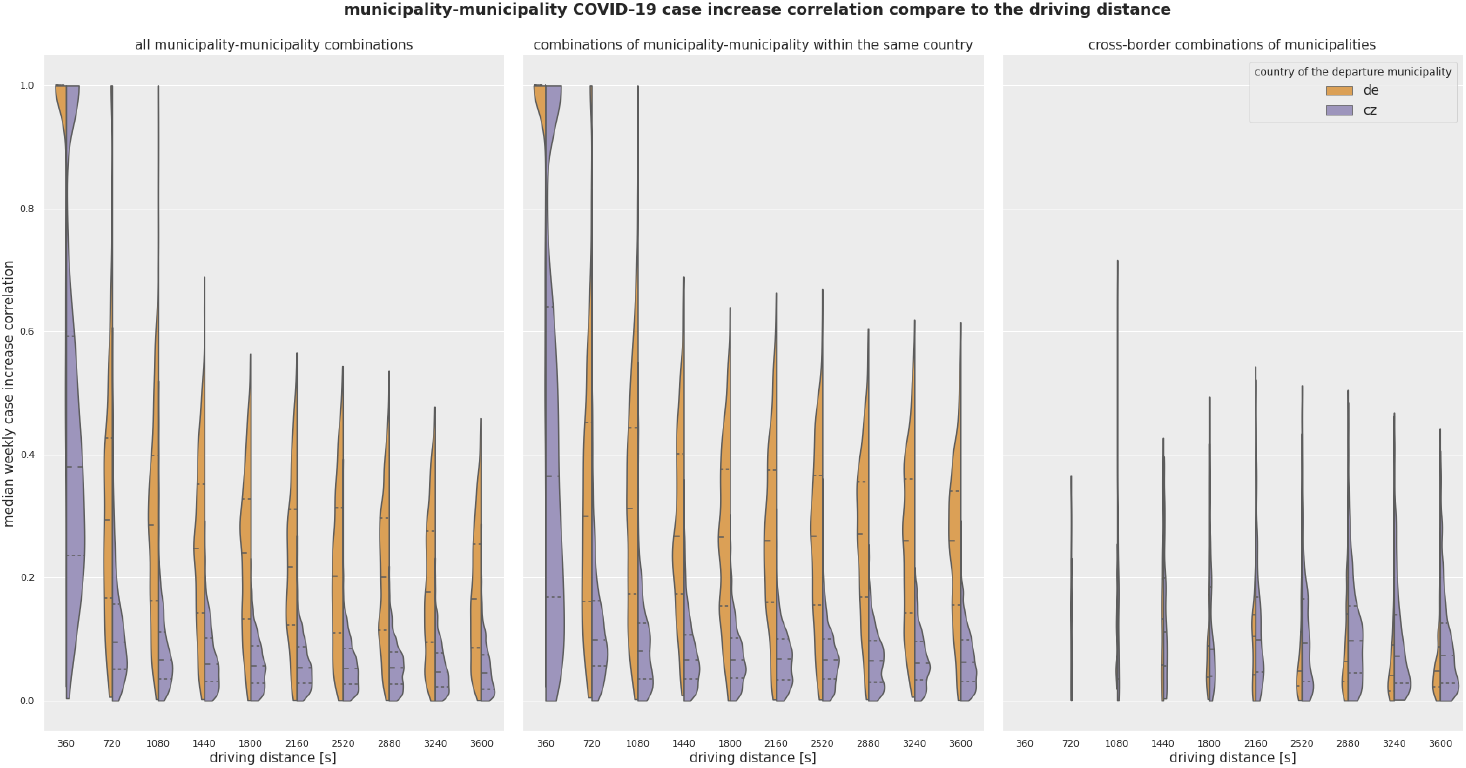
Spatial autocorrelation graph. The dependence of the municipality-municipality correlations in the case numbers on the temporal distance. The chart highlights the similarity between two municipalities (quantified by the correlation number) on their temporal distance. We notice a strong similarity of case numbers across municipalities within the same country. Such correlations tend to be significant when the municipalities are within 20 minutes of each other, implying strong spatial autocorrelation. A similar relationship of autocorrelation in case numbers at short temporal distances is not present for pairs of municipalities separated by the national border. This finding suggests a strong effect of the national border on the spread of the virus.

**Figure S4.**
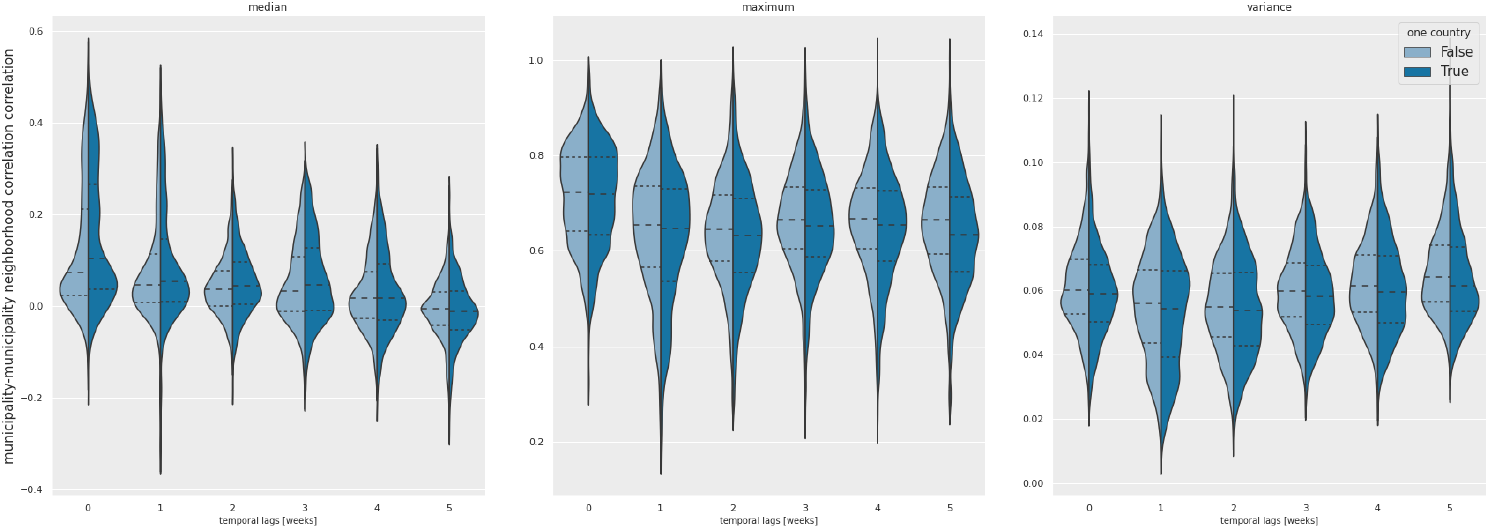
Correlation values of the municipality-municipality temporal trends considering the neighborhood defined by one hour of driving distance. The dark blue color stands for all combinations of municipalities in one country, the light blue color for all municipality-municipality combinations. While the median correlation values change little across various time lags, we note a difference in the distribution of values within the higher part of the scale(75th percentile). This refers to the situations when not all municipalities have similar trends in case numbers, but the correlation is present much more often when the temporal lag is lower. The values are also notably higher when considering only the combination of municipalities within one country and the lag of zero or one week. Maximum values of the correlation for each municipality show a significant decrease in the time lag of one week, while all the other time lags retain similar distributions. More interesting is the comparison of the variance in the neighborhood municipality-municipality correlations. In this case, the time lags of one and two weeks have the lowest numbers; slightly higher numbers are reached when we consider the municipalities on the other side of the border. This analysis may indicate overall spatio-temporal trends of the virus spread. While the median values do not change significantly over time, the distributions of the medians and the values of the maximum correlations reveal that the similarity within the neighborhood is higher when there is no time lag. In contrast, the variance analysis revealed the highest consistency in correlation values when the time lag is one or two weeks. Additionally, correlations are generally higher when we consider only municipalities from one country, which again highlights the effect of the border.

**Figure S5.**
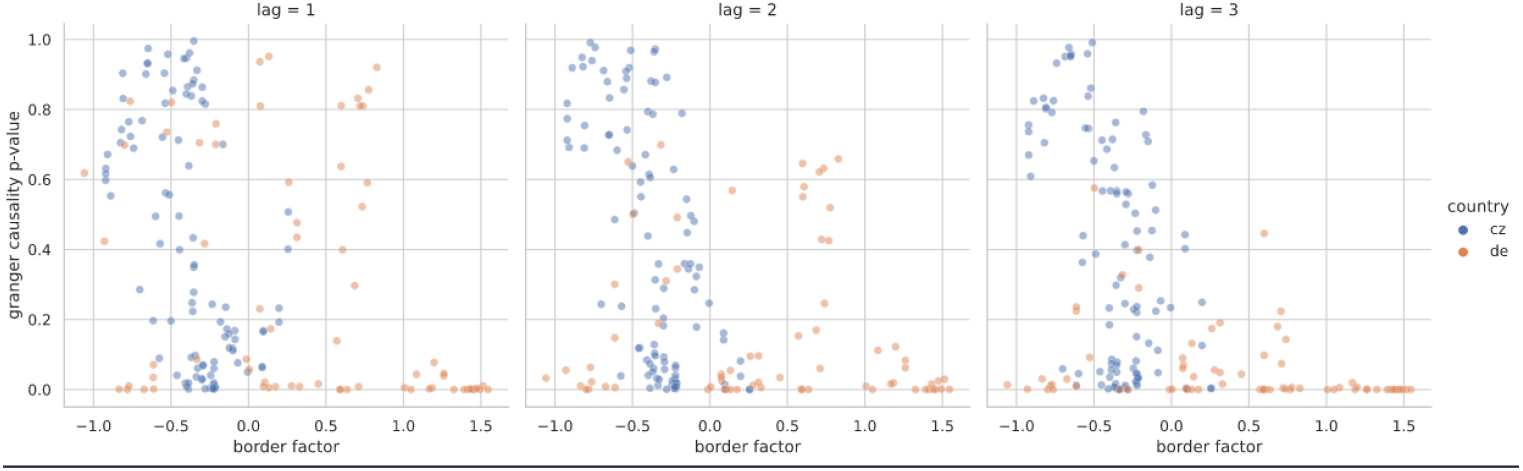
The comparison of the weighted border factor calculated by the beta regression model and the granger causality test p-value considering the temporal lag of 1, 2, and 3 weeks. The granger causality values were calculated by comparing the case values in the neighboring municipalities in the same country to the case values in neighboring municipalities on the other side of the border.

## Notes

### Competing Interest Statement

The authors have declared no competing interest.

### Summary of Updates

The authors' affiliations have been corrected.

